# Monitoring Emergency Medical Services Frequent Use: Protocol for a Mixed-Methods Study Using Routinely-Collected Data

**DOI:** 10.64898/2026.09.07.26362041

**Authors:** Laura Maruster, Durk-Jouke van der Zee, Bert Dercksen

## Abstract

**Introduction:** Aging populations are placing increasing pressure on Emergency Medical Services (EMS) providers to address the potentially suboptimal use of scarce resources. EMS utilization that falls outside the established deployment framework, referred to here as ‘non-indicated use’, may constitute an important source of such suboptimal resource use. Frequent users of EMS represent a key target group in efforts to reduce non-indicated utilization. The focus on frequent users is driven by the substantial workload and costs associated with their use of EMS, as well as by the need to address potential underlying quality-of-care issues reflected in high, persistent, or intensive EMS utilization.

Addressing possible non-indicated EMS use among frequent users requires the development of monitoring systems that can be applied in both clinical practice and research to identify such use and support appropriate action once it has been detected. This study aims to explore the relationships between the drivers of frequent EMS use, potential remedies for non-indicated use, and subsequent reductions in utilization. Based on these insights, the study seeks to develop, through prototyping, a framework to guide the design of monitoring systems for research and clinical practice.

**Methods and analysis:** We will employ a mixed-methods observational study design. A systematic literature review and interviews with stakeholders who play key roles in the EMS referral process will be conducted to identify the drivers of frequent use and ‘non-indicated’ EMS use and their potential remedies. Pseudonymized routinely collected data covering 2013-2026 from three Dutch EMS providers will be analyzed using machine learning (ML) and artificial Intelligence (AI) techniques to develop a typology of drivers and trustworthy, interpretable models for identifying frequent use and non-indicated use.

A prototype monitoring system will be developed to demonstrate the online and the offline functionalities of the proposed framework for use in both clinical practice and research. The prototype’s usability, trustworthiness, and interpretability will be evaluated in collaboration with relevant stakeholders involved in the EMS referral process.

**Ethics and dissemination:** The data is routinely collected for administrative purposes only. Accordingly, the data will be completely pseudonymized, and as such this study falls outside the scope of the Dutch Medical Research Involving Human Subjects Act. After assessment we obtained a full waiver for using pseudonymized data from the EMS services from the Medical Ethics Review Board of the University Medical Center Groningen, The Netherlands. Patients and/or the public are not involved in the design, conduct, reporting, or dissemination plans of this research. Interviewees and participants in evaluating prototype monitoring systems (e.g. EMS clinicians, medical directors, ambulance nurses, emergency physicians, general practitioners (GP’s), policy makers and police officers) will provide informed verbal consent prior to their participation in the study. The study findings will be disseminated through peer-reviewed publications, presentations at conferences, and in social media. Also, summary reports will be made available to participating institutions and relevant stakeholders.

**Strengths and limitations of this study:**

- The study will provide comprehensive insights into underlying drivers of frequent and ‘non-indicated’ EMS use (i.e. EMS care utilization that falls outside the agreed upon deployment framework), potential remedies, and the resulting implications for patients, EMS providers, and other healthcare providers.
- A framework for the systematic design of EMS monitoring systems will be developed to identify and support action on frequent and non-indicated EMS use. The framework is firmly grounded in both the scientific literature and clinical practice.
- An AI-based approach using routinely collected EMS records will be proposed to model frequent and non-indicated EMS use, that fosters trustworthiness and interpretability for relevant stakeholders.
- The prototype monitoring system will demonstrate the on-line and off-line functionalities of the proposed framework and evaluates their usability, trustworthiness, interpretability, and relevance for clinical practice.
- The use of routinely collected EMS data from three provinces of Northern Netherlands operating within the same geographical area and healthcare system may limit the generalizability of the findings.

## INTRODUCTION

In many countries, Emergency Medical Services (EMS) are under increasing pressure due to rising demand for services ^1–4^ and staff shortages^5^, with population aging being an important contributing factor. Ensuring the optimal use of EMS personnel and resources for providing pre-hospital patient triage, treatment and transport is therefore paramount. This creates a need to monitor current patterns of resource use, identify and assess potential instances of suboptimal utilization, and take appropriate action when such instances are detected. An improved understanding of suboptimal EMS use should include the utilization of EMS care that falls outside the agreed deployment framework of EMS – referred to here as *non-indicated use* – and that could be more appropriately addressed by other care providers (if there is a need). For instance, a patient with a urinary tract infection who presents with severe abdominal pain may require antibiotic treatment. In the Dutch healthcare system, such a patient would ideally be treated by a general practitioner, who is authorized to prescribe antibiotics. In such cases, EMS utilization should preferably be avoided, as paramedics are not authorized to prescribe antibiotic treatment

### Background and rationale

Frequent users of EMS are considered a key target group in efforts to reduce potentially suboptimal use of health resources. Between 0.2% and 23% of patients served by EMS are considered frequent users, accounting for approximately 1.4% to 40% of total ambulance utilization^6^. Much of this variation can be explained by the different thresholds used in determine frequent use, ranging from fewer than one transports per year to several dozen transports per year, with reported averages generally range between three and ten EMS responses per year^7,8^. The focus on frequent users is readily explained by the substantial workload and associated costs attributable to their EMS use, as well as by the need to address possible underlying quality-of-care issues. Such issues may be reflected in temporal patterns of EMS utilization, with the (high) number, persistence, and intensity of calls serving as crude indicators of potentially problematic use^6–9^.

Several studies have examined how patient-related characteristics, such as age, gender, mobility limitations, social economic status, mental or physical health conditions, chronic disease, comorbidity, and alcohol or drug misuse are associated with EMS frequent use^12–26, 28^. To address potentially suboptimal use of EMS resources, various interventions have been proposed and implemented, including care plans, changes in policies and care pathways, and enhanced triage practices, with the aim of reducing call frequency and unnecessary ambulance transports.^1,6,10–13^

Non-patient-related drivers of frequent EMS use should also be taken into account. This may include characteristics of healthcare network logistics, such as the availability and accessibility of primary care^18^, treatment-related factors, such as radiotherapy requiring patients to undergo a series of treatment sessions, and the organization of regional healthcare systems, such as hospital specialization creating a need for interhospital transports^27^. Importantly, the nature of these underlying drivers may determine the scope and type of interventions required to reduce frequent EMS use. For example, advances in radiotherapy may enable shorter treatment series length and thereby reduce transport requirements^27^. Similarly, reducing interhospital transports may require regional optimization of hospital resource allocation and referral arrangements, involving trade-offs among the interest and priorities of different stakeholders^27^.

Although the literature identifies relationships between different categories of drivers and frequent EMS use, inappropriate frequent use remains insufficiently defined. To facilitate the translation of research insights into EMS referral decision-making, the identified drivers need to be related to the portfolio of services that EMS is expected and authorized to provide. As a first step, we propose to distinguish between ‘indicated EMS use’ - EMS use that is clinically justified, and ‘non-indicated EMS use’ - EMS use that falls outside the agreed deployment framework. In the Netherlands, this deployment framework is defined by formal national protocols that specify treatment options for ambulance personnel and aim to support the provision of optimal ambulance care within applicable legal limits and the professional competencies associated with the training and qualifications of ambulance personnel^29^. These protocols serve as guidelines for ambulance care, while allowing ambulance personnel to deviate from these in specific circumstances when such deviations can be adequately justified. The national protocols cover the full EMS portfolio of services, ranging from immediate transport to a hospital (“scoop and run”) to extensive advanced life support interventions at the scene site (“stay and play”). The protocols are continuously evaluated and, when necessary, revised by a national committee^29^.

To further clarify the concept of ‘non-indicated EMS use’, two examples involving patients with mental health conditions are provided. As an example of ‘non-indicated’ transport, consider an institutionalized patient with mental disorder who leaves the institution without permission and travels 300 km. If an incident involving the patient subsequently occurs, existing protocols may require an ambulance to transport the patient back to the institution (potentially even against patient’s will). Such a transport places a considerable burden on EMS resources and occupies an ambulance that could otherwise be available for an acute emergency call. A potential remedy would be a dedicated, potentially commercial, transport service staffed by personnel specially trained to transport such patients back to the institution. By contrast, an example of ‘indicated’ EMS use is a patient experiencing an acute psychotic episode who is found in a public place, requires urgent assessment and antipsychotic treatment, and needs immediate transport home to an appropriate care setting.

Although the literature acknowledges EMS frequent use and proposes interventions aimed at reducing such use, it provides limited guidance on how potentially ‘non-indicated’ frequent EMS use can be systematically identified and addressed in clinical practice. Consequently, there is also little evidence on the design and implementation of monitoring systems specifically targeting frequent EMS use. One of the few reported examples mentions a rudimentary “within-service” flagging system intended to discourage repeated EMS calls^12^. Addressing this gap is essential for advancing the field and ensuring that research findings are translated into meaningful improvements in clinical practice and appropriate use of EMS resources. To this end, methodological and tool-based support is needed to enable the systematic identification, assessment and management of potentially ‘non-indicated’ frequent EMS user. Such monitoring systems should build on insights derived from EMS practice and make effective use of routinely collected data generated by dispatch-center personnel, ambulance nurses, medical specialists, and other healthcare providers. By integrating these sources of information, monitoring systems could support targeted interventions and contribute to a more efficient use of emergency care resources. Ultimately, EMS monitoring systems should also support the regular evaluation of the scope of EMS services, operational practiced and, and deployment protocols.

One of the main challenges in monitoring frequent EMS use in clinical practice is the limited availability of routinely collected data targeting frequent use. In most settings, such dedicated data are not available. However, routinely collected EMS records may contain relevant information that can support the identification and analysis of frequent use^6^. For instance, a recent study using EMS records for the province of Drenthe, the Netherlands analyzed free-text clinical narratives to support the identification of older patients at risk of frequent EMS use^41^. The study demonstrated that unlocking information contained in routinely collected EMS data, including triage reports written in natural language, poses considerable challenges because of complexity and variability of such data. By applying machine learning and advanced artificial intelligence techniques to analyze and interpret these heterogenous healthcare data, the study showed that the identification of frequent EMS users could be improved. Similar findings on the use of ML and AI techniques to extract clinically relevant insights from routinely collected healthcare data have been reported in the broader healthcare literature, see for instance^30^.

### Objectives

This study investigates the drivers of frequent EMS use and examines how non-indicated frequent can be reduced. More precisely, the study explores the relationship between the drivers of frequent EMS use, possible remedies and subsequent reductions for non-indicated EMS use. Building on these insights, the study proposes a framework for designing monitoring systems for frequent EMS use. The framework combines advanced AI-based methods with a user-centered approach to foster trustworthiness and interpretability for all relevant stakeholders.

The research objectives are:

1. Define frequent EMS use by distinguishing between indicated and non-indicated use, thereby supporting the identification of potentially avoidable EMS use in the future.
2. For selected patient groups, identify patient- and non-patient related drivers of frequent and non-indicated EMS use, assess their impact on EMS resources, and explore potential remedies to address these drivers.
3. Develop and evaluate a framework for systematic monitoring of frequent EMS use in clinical practice, with the aim of reducing non-indicated EMS use. A prototype monitoring system will demonstrate the core functionalities of the proposed framework and will be evaluated through use cases in selected Northern Netherlands EMS organizations.

## METHODS AND ANALYSIS

This study will combine quantitative analyses of routinely collected pre-hospital data with qualitative evidence obtained through stakeholder interviews. The proposed study will use a mixed methods design approach and will be conducted and reported in accordance with RECORD^31^ (REporting of studies Conducted using Observational Routinely-collected health Data) checklist and TRIPOD+AI^32^ (Transparent Reporting of a Multivariable Prediction Model for Individual Prognosis, and Diagnosis+AI Checklist for Prediction Model Development and Evaluation).

### Study setting

The study will be conducted in the provinces of Fryslân, Groningen and Drenthe, the Netherlands, which together have a total population of 1,774,019, and a population density of 208 inhabitants per square kilometer^33^. In Northern Netherlands there are twelve hospitals currently, providing different levels of care, including tertiary care. Patients in the Northern Netherlands predominantly rely on hospitals located within the region, although some receive care outside the region because they live near its borders or require specialized care.

EMS care in the Northern Netherlands is provided by three EMS organizations, each serving one province and operating through a combined network of 51 ambulance bases^34^. Together, these organizations carry out more than 120,000 patient transports annually. Their services include urgent treatment and transport, planned transports to hospitals, and planned transports to other care providers, particularly nursing and care homes. Dedicated ambulance fleets are used for different types of transport. The Advanced Life Support (ALS) fleet is primarily deployed for urgent care and is subject to high staffing, equipment and competence requirements, enabling the treatment and transport of critically ill patients. Basic Life Support (BLS) fleet generally involves lower staffing and resources. More recently, Medium Complex (MC) ambulance care has been introduced in the Netherlands. This category encompasses ambulance care that exceeds basic planned transport but does not require the full capabilities of ALS care, thereby providing an intermediate level of care between planned transport and advanced ambulance (ALS) care^35^. Depending on patients’ clinical needs, planned transport may require ALS, MC or BLS staff and resources. When those needs are limited, independent travel or taxi transport may be sufficient. All EMS transports across the three provinces are coordinated through a single dispatch center serving all the entire region.

#### EMS in the Netherlands – staffing, protocols and referral process

Emergency medical care in the Netherlands is provided by specialised ambulance nurses or bachelor’s-degree-qualified paramedics, working alongside specialised ambulance drivers^36^.

EMS care is highly standardized and guided by national protocols. EMS professionals across the country are required to adhere to more than 70 protocols^29, 36^. To maintain high standards of care, EMS staff participate in continuous medical education and training after their initial professional qualification. During a deployment, the ambulance team operates with a high degree of professional autonomy. However, a supervising physician is available for consultation by phone 24 hours a day. Following a deployment, the quality of care is assessed by the supervising physician through, among other methods, a review of the Electronic Deployment Report^36^.

EMS care is coordinated through regional dispatch centers. Ambulance care may be initiated by members of the public via the national emergency number 112, directly by healthcare professionals such as hospital physicians and GP’s, or by other authorities, including the police. All calls are handled by EMS dispatchers who are specialised nurses and use a digital triage system to determine the most appropriate response. Depending on the situation, the response may involve dispatching one or more emergency units, such as ambulances or helicopters, referring the caller to another healthcare provider, or providing self-care advice. In addition to emergency deployment, ambulances are also used for less urgent or non-urgent transports. These transports are often planned in advance, for example for dialysis, radiotherapy, or scheduled hospital appointments, and are generally initiated by healthcare professionals.

### Patient and public involvement

Patients and/or the public will not be involved in the design, conduct, reporting, or dissemination of this research. To evaluate the study results and develop use cases, we will involve representatives of different stakeholder groups, including general practitioners, EMS dispatch-center personnel and nurses, medical specialists, medical experts and researchers.

### Study design

The proposed research will adopt a mixed-methods, retrospective observational design. Pseudonymized routinely collected EMS records, comprising both structured and free-text data will be analyzed quantitatively to identify drivers and remedies of frequent and non-indicated EMS use, and support the development of the monitoring framework and prototype system. Stakeholder interviews will provide complementary qualitative insights into the drivers of frequent and non-indicated EMS use and possible remedies. Figure 1 visualizes the study design, which is described below.

**Figure 1.**
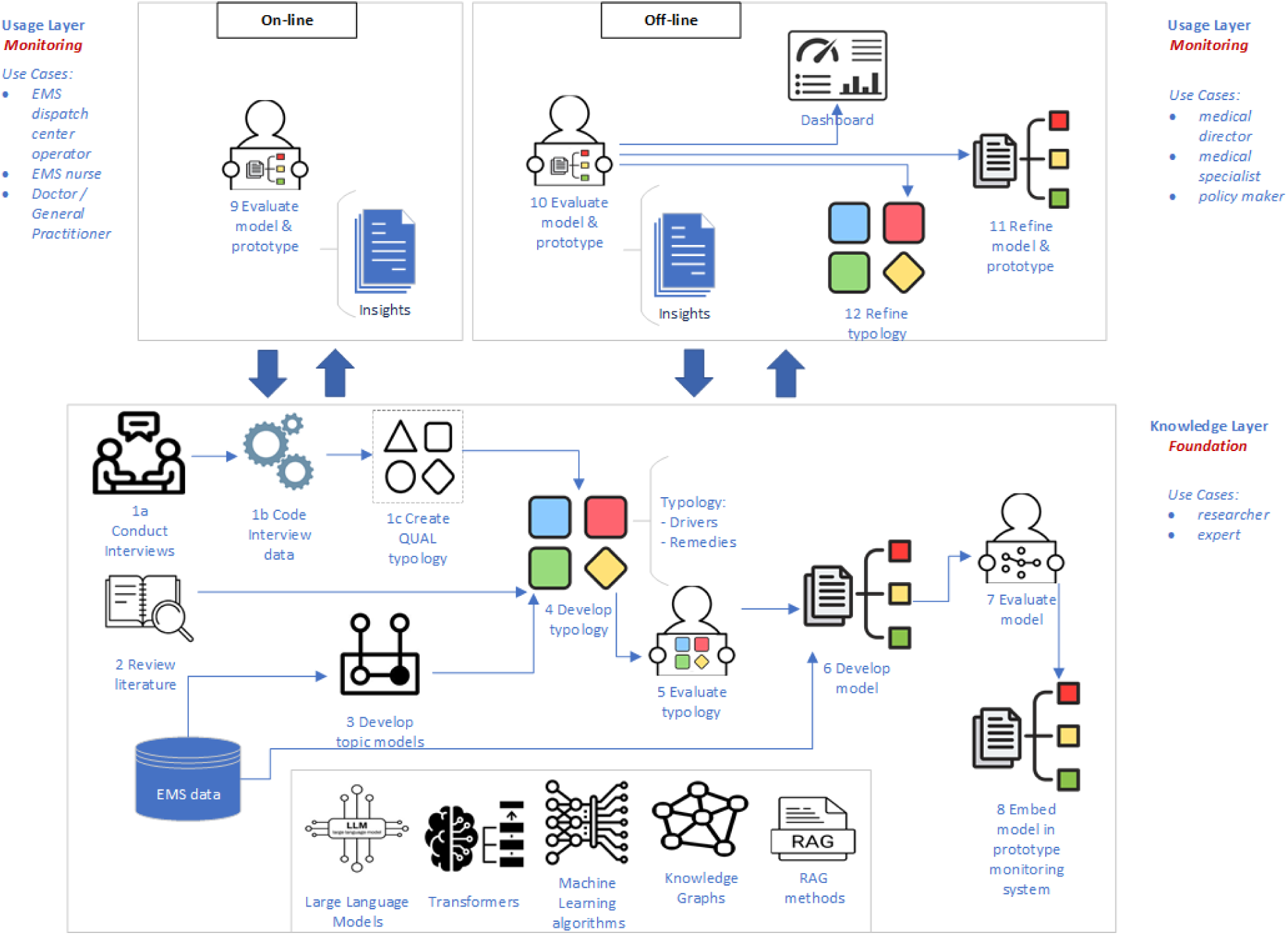
Study design for EMS monitoring system design and evaluation.

By contrast, an example of ‘indicated’ EMS use is a patient experiencing an acute psychotic episode who is found in a public place, required urgent assessment and antipsychotic treatment, and needs immediate transport home to an appropriate care setting.

The study has two main aims, corresponding to a **Knowledge**, and a **Usage** layer:

#### Aim 1 - Knowledge layer: identifying the drivers of EMS frequent and ‘non-indicated’ EMS use, and their remedies

To develop the Knowledge layer (Figure 1), we will employ a mixed-methods setup. Firstly, a systematic literature review will be conducted to identify the drivers of frequent EMS use, distinguish between ‘indicated’ and ‘non-indicated’ use, and identify potential remedies. The review will be reported in accordance with the PRISMA guidelines for systematic reviews. Secondly, the drivers of frequent use, ‘non-indicated’ use, as well as the potential remedies identified from literature will inform the development of the interviewing protocol. Interviews will be conducted to validate and deepen the findings from the literature, thereby supporting the development of an initial typology of drivers and remedies. Thirdly, pseudonymized routinely collected EMS records covering all patient transports carried out by EMS providers for the Northern Netherlands will be analyzed to challenge and refine the initial typology of drivers and remedies.

The typology of drivers and their corresponding remedies, together with classification models to classify indicated and non-indicated EMS use will be embedded in a prototype monitoring system. The prototype will integrate the activities from the Usage layer (see the upper part of Figure 1). For example, a classification model for the driver “dependent on regular clinical intervention” could include a decision rule such as: “If a patient has a chronic condition requiring regular clinical intervention, then EMS transport is justified”, and thereby is assigned to class ‘indicated EMS use’. Examples include patients who require regular dialysis, chemotherapy, or radiotherapy.

#### Aim 2 - Usage layer: Use cases for monitoring EMS use

The Usage layer comprises the monitoring activities performed in clinical practice. Both *off-line* and *on-line* use of the monitoring system are acknowledged in practice.

*On-line usage* implies real-time interaction with the system and supports day-to-day decision-making of EMS dispatch center operators, ambulance nurses, medical specialists, and GP’s. For example, an EMS dispatcher may use the monitoring system alongside a digital triage system to determine the most appropriate response to an incoming call. Typically, these users may query the system to distinguish between frequent and non-frequent EMS users, retrieve patient characteristics, assess risks, and predict the most appropriate type of intervention, such as transport to hospital versus treatment at home. The system may also support decision regarding the most appropriate type of vehicle and recommend interventions based on patient characteristics. In addition, it aims to suggest suitable remedies tailored for the specific situation at hand.

*Off-line usage* involves monitoring activities aimed at the continuous development, maintenance, evaluation, and improvement of the monitoring system and its components. These activities include examining patterns of frequent use, identifying (new) patient groups, and conducting longitudinal analysis of (persistent) frequent users. The findings may inform revisions to protocols, adjustments to individual patient care plans, and the development of care pathways tailored to specific patient-groups. The expected users of the offline functionalities include medical specialists, medical directors, policymakers, experts and researchers. Their activities involve the periodic review and evaluation of the system’s functionalities and outputs, including the typology of drivers and remedies, machine learning models, and classification criteria for frequent user and non-indicated EMS use. Such reviews also support the detection of changes in patterns, drivers and patient characteristics over time. In addition, the system is intended to support patient-level and patient-group-level analysis, such as identifying groups for whom targeted actions, or specific interventions may be required.

To develop, test and refine the prototype monitoring system, use cases will be designed for both on-line and off-line modes of use. These use cases will involve all relevant stakeholder groups, including EMS dispatch centre operators, ambulance nurses, GP’s, medical specialists, medical directors, and policymakers. Through these use cases, the prototype monitoring system developed under Aim 1 will be tested for usability, understandability and trustworthiness. The thick arrows in Figure 1 between the Usage and the Knowledge layers clarify a feedback loop between the two layers: the typology of drivers and the classification models developed in the Knowledge layer are used to support on-line and off-line monitoring activities. Conversely, insights coming through the on-line and off-line use cases are analyzed and incorporated into the relevant components of the Knowledge layer, allowing the system and its underlying knowledge models to be progressively refined. For example, an initial decision rule stating “if a patient presents symptom A, then perform intervention B” may be revised on the basis of insights from system use to “if a patient presents symptom A, then perform intervention C instead of intervention B”. The revised rule is subsequently incorporated into the Knowledge model within the Knowledge layer.

### Data sources and data collection procedures

The study will integrate two types of data: qualitative interviews with stakeholders, and prehospital routinely collected data.

#### Interviews

Primary data will be collected through semi-structured interviews with knowledgeable stakeholders involved in the EMS referral process, including EMS medical directors, ambulance nurses, emergency physicians, EMS dispatch center personnel, GP’s, and non-medical professionals such as police officers. The interviews will explore the drivers of frequent use and ‘non-indicated’ use, as well as potential remedies. The interviews will be guided by interview protocols tailored to the different stakeholder groups. All interviews will be audio-recorded, transcribed and systematically analysed.

#### Prehospital data – EMS records

Pseudonymized retrospective routinely collected prehospital data will be provided by three EMS providers for the Northern Netherlands. The data will comprise selected fields from EMS records covering all patient transports carried out by these EMS providers for the Northern Netherlands, between January 1, 2013 and December 31, 2026. Each EMS call is documented according to a standardized dispatch protocol and includes timestamps, information on patient triage, vehicle deployment and resource use. In addition to structured fields, such as patient age, medication use, and transportation date and time, the EMS records contain free-text fields describing patient triage, clinical circumstances, and logistics aspects. The free-text data are recorded by personnel at the regional EMS dispatch centre and by ambulance nurses attending the patient at the scene.

The data will be cleaned to identify and remove noise. For example, records referring to transport from or to a non-existent location or postal code will be considered noise and excluded from the analysis. Missing data are an inherent characteristic of routinely collected data and will not be assumed to be missing at random. Therefore, no imputation or exclusion strategies will be applied, as these approaches may introduce bias. Instead, missing data will be preserved and analyzed descriptively.

Routine data is pseudonymized locally. Quasi-identifiers will be securely transferred to a trusted third party for privacy-preserving record linkage. The linked and de-identified research dataset will be analysed exclusively within a secure research environment.

### Data analysis

#### Analysis of interview data

The qualitative data obtained from interviews will be analyzed using qualitative content analysis, combining thematic and theoretical coding. For example, previously identified categories of drivers such as socioeconomic status, mental health problems, physical health problems, chronic disease, medical comorbidity, and alcohol or drug misuse^37–39^, will be used as initial themes to guide the thematic coding process. To enhance coding reliability, the interview data will be independently coded by two researchers. The drivers of frequent EMS use and ‘non-indicated use’, together with their associated remedies, identified through the systematic literature review (see step 2 in Figure 1) will be compared and integrated with the interview findings. The combined findings will be organized into a preliminary qualitative typology (Figure 1; steps 1a, 1b and 1c). For example, for the driver “mental health problem”, both the literature review and the interviews may identify specific conditions, such as “psychotic” or “neurocognitive” disorders. Whether ambulance dispatch is ‘indicated’ or ‘non-indicated’ will depend on the patient’s specific condition. For example, an aggressive psychotic patient who is unable to communicate may require sedation, making ambulance transport necessary, and therefore indicated.

#### Analysis of EMS routinely collected data

The pseudonymized EMS data will be analyzed with both unsupervised and supervised machine learning and artificial intelligence methods.

#### Unsupervised learning methods

The preliminary qualitative typology of drivers and remedies for non-indicated use (step 1c in Fig.1) will be refined further using the routinely collected data. Topic models will be developed to automatically identify latent themes and patters associated with ‘non-indicated’ EMS use, its underlying drivers, and potential remedies (step 3 in Fig.1). Topic modelling is a Natural Language Processing (NLP) approach used to uncover the latent thematic structures within large collections of textual data^40^. The topic model results (step 3 in Fig. 1) will be then compared with the preliminary (qualitative) typology (step 1c in Fig.1), which will result in an enriched typology of drivers, types of non-indicated EMS use and their associated remedies (step 4 in Fig.1). Finally, the enriched typology will be evaluated by a panel of EMS professionals involved in the referral process, including medical directors, ambulance nurses, emergency physicians, EMS dispatch center personnel, and general practitioners (step 5 in Fig. 1).

#### Supervised learning methods

The validated typology (step 5 in Fig. 1) including the identified classes of drivers of frequent use and ‘non-indicated’ EMS use, will be employed to develop classification models. The development of classification models will involve iterative training, validation, and testing to obtain predictive models with high accuracy and performance (step 6 in fig. 1). The resulting models will classify EMS transports as either ‘indicated’, or ‘non-indicated’ on the basis of the underlying driver, such as ‘abuse’, ‘misuse’, social-related factors, etc. For instance, chronic conditions such as kidney failure requiring dialysis and Chronic Obstructive Pulmonary Disease (COPD) may contribute to frequent EMS use. In the case of kidney failure, planned ambulance transport to a dialysis center constitutes ‘indicated’ use. By contrast, patients with COPD who do not adhere to prescribed medication may experience exacerbations that can lead to emergency hospitalisation and ambulance transport. Although the ambulance transport may be clinically indicated at the time of the acute exacerbation, the broader pattern of repeated EMS use may be considered potentially preventable and thus ‘non-indicated’.

To develop the classification models (step 6 in Fig. 1), machine learning algorithms capable of handling heterogeneous data types, such as Random forest, Gradient Boosting, XGBoost, and LightGBM will be employed. The available data comprise both structured data, such as age, medication use, medical specialty, and transport type, and free-text data documenting the clinical observations, and circumstances at the scene. To support their use in clinical practice by ambulance professionals, the developed classification models must be trustworthy, interpretable and transparent. Their clinical relevance will be evaluated by an expert panel (step 7 in Fig. 1). Classification models demonstrating satisfactory predictive performance and clinically acceptability will be selected for further development and integration into the prototype monitoring system (step 8 in Fig.1). Predictive performance will be assessed using appropriate measures such as accuracy, precision, recall, and the F1-score.

#### Natural Language Processing and AI methods

The topic-modelling analysis (step 3 in fig. 1) will be conducted using NLP methods such as BERTopic^40^. Given the highly confidential natura of data, locally deployed LLM’s will be used. Retrieval-Augmented Generation (RAG) methods will be employed to enhance LLM output by providing access to relevant external knowledge^42^. To support the explainability of the classification models, knowledge graphs will be developed to provide structured representation of relevant entities, and their relationships^43^. These representations may help contextualise model predictions and make relationships between drivers, types of EMS use, and potential remedies more transparent to users. Because the development and deployment of LLM-based models are computationally demanding, model development will be hosted on the High-Performance Computing (HPC) cluster of the University of Groningen, The Netherlands.

#### Development and evaluation of the prototype monitoring system with use cases

Finally, a prototype monitoring system will be developed and validated through use cases to demonstrate the core functionalities of the proposed framework. The prototype will be evaluated in terms of usability, understandability and trustworthiness and will refined based on use cases involving relevant stakeholder groups (EMS dispatch centre operators, ambulance nurses, GP’s, medical specialists, medical directors, and policy makers) (steps 9, 10, 11, 12 in Fig.1). The scope of this study is limited to the development and evaluation of the prototype monitoring system, and does not include the full-scale development, deployment, or implementation of an operational monitoring system. Instead, the study will provide a validated blueprint that can guide the future development and implementation of such systems in clinical practice.

## ETHICS AND DISSEMINATION

The data in this study are routinely collected for administrative purposes only. The research dataset will be fully pseudonymized before being made available to the research team. Accordingly, the use of this data falls outside the scope of the Dutch Medical Research Involving Human Subjects Act. Following assessment by the Medical Ethics Review Board of the University of Groningen, a full waiver was granted for the use of pseudonymized EMS data (reference number METc 2022/053). Patients and members of the public will not be involved in the design, conduct, reporting, or dissemination of this research. Interviewees and participants in the evaluation of the prototype monitoring systems, including EMS clinicians, medical directors, ambulance nurses, emergency physicians, general practitioners, policy makers and police officers will provide informed consent before participating in the study. The study findings will be disseminated through peer-reviewed publications, presentations at conferences, and appropriate social media channels. Also, summary reports will be made available to participating institutions and relevant stakeholders.

### Risk-benefit assessment

In many countries, Emergency Medical Services (EMS) providers are under increasing pressure to address the potentially suboptimal use of their scarce resources, as aging populations challenge both the demand and the supply of these resources. The outcomes of this study will demonstrate the value of systematically monitoring and regularly scrutinizing the scope, activities, and protocols of EMS in order to help prevent ‘non-indicated’ EMS use, with frequent users serving as a key focal group.

The study will propose an actionable framework for development and use of monitoring systems aimed at preventing frequent non-indicated EMS use, an area in which such approaches are currently lacking or, at best, fragmented^12^. The framework relies on a stepwise approach and recommends the use of analysis methods, including AI-driven methods. The use of framework will be demonstrated and evaluated through the development of a prototype monitoring system that supports two complementary modes of use.

Firstly, the prototype monitoring system will support on-line use in daily clinical practice by facilitating better-informed referral and dispatch decisions, thereby directly contributing to the reduction of non-indicated EMS use. Secondly, its off-line use will facilitate continuous learning from current clinical practice by enabling the integration of new insights and the adaptation of the system’s functioning over time. This capacity ongoing refinement will help ensure that the monitoring system remains responsive to changes in user needs, EMS regulations, patient characteristics, health services and systems and other related developments.

The development of monitoring systems such as the one envisioned in this study is likely to benefit substantially from the use of routinely collected data and AI-driven analytical methods. While the reuse of EMS records increases the efficiency of data collection, AI-driven methods enable the efficient and effective analysis of large volumes of data, including unstructured free text clinical narratives^30^. Early evidence of this potential is provided by a recent related study involving EMS records from the province of Drenthe, the Netherlands, in which free-text clinical narratives were analyzed to support the identification of older patients at risk of becoming frequent EMS users^41^.

By developing monitoring systems, this study will also advance the knowledge of the drivers of frequent use and non-indicated (avoidable) EMS use, and their possible remedies, by examining and scrutinizing the full range of services provided by EMS within the broader acute care network. Accordingly, the understanding of these drivers and their associated remedies is expected to become broader and more systematically elaborated than has been portrayed in the literature to date. Apart from its direct relevance to clinical practice, this broader perspective will help identify gaps in current knowledge and opportunities for future research in the field.

Although regional characteristics of the study setting may introduce some bias, resulting in differences in the types and relative importance of drivers of frequent EMS use and related remedies, such variation is not expected to compromise the overall structure of the proposed design approach for EMS monitoring system or limit its broader applicability.

## Data Availability

The raw third-party data utilized in this study are proprietary to UMCG Ambulance Care Tynaarlo, Groningen and are subject to licensing restrictions. Consequently, the authors are not legally permitted to distribute the raw data. Access requests can be directed to. However, the authors' custom analysis scripts, code, and de-identified derived data layers used to generate the findings of this prospective study will be openly available in a GitHub repository.

## Acknowledgments

We acknowledge that chatbot ChatGPT Plus (OpenAI) was used solely to improve the clarity and grammar of selected passages of the manuscript. The tool was not used to generate the research questions, study design, data analysis, interpretation of results, or scientific conclusions. All AI-assisted revisions were critically reviewed and, where necessary, modified by the authors, who take full responsibility for the final content of the manuscript.

## Authors contribution

LM, DJZ and BD conceived the study idea. LM and DJZ designed the study and elaborated the methodology. All authors will support data collection, analysis and implementation of the study. LM drafted the manuscript and all authors contributed to the edit of the manuscript. All authors read and approved the final version.

## Data availability statement

No additional data are available.

## Funding statement

This research received no specific grant from any funding agency in the public, commercial or not-for-profit sectors.

## Conflicts of interests

All authors have completed the ICMJE uniform disclosure form at http://www.icmje.org/disclosure-of-interest/ and declare: no support from any organisation for the submitted work; no financial relationships with any organisations that might have an interest in the submitted work in the previous three years; no other relationships or activities that could appear to have influenced the submitted work.

## Notes

### Competing Interest Statement

The authors have declared no competing interest.

### Author Declarations

The Medical Ethics Review Board of the University Medical Center Groningen waived ethical approval of this work (reference number METc 2022/053).

